# A vagal influence on schizophrenia? A nationwide retrospective cohort of vagotomized individuals

**DOI:** 10.1101/2024.01.30.24301418

**Authors:** Cornelia F Richter, Karolina P Skibicka, Urs Meyer, Sabine Rohrmann, Jean-Philippe Krieger

## Abstract

**Background and Objectives:** Emerging preclinical evidence suggests that vagal signals contribute to the development of schizophrenia-related abnormalities in brain and behavior. Whether vagal communication in general, and its impairment in particular, is a risk factor for schizophrenia in humans remains, however, unclear. Vagotomy, the surgical lesion of the vagus nerve, was routinely performed as a treatment for peptic ulcer before modern treatment options were available. Hence, the primary aim of this study was to investigate whether vagotomy modulates the subsequent risk of developing schizophrenia. Moreover, given the existence of diverse vagotomy techniques (i.e., “truncal” or “selective”), our secondary goal was to test whether the extent of denervation modulates the risk of schizophrenia.

**Methods:** Using a nationwide retrospective matched cohort design, we identified 8,315 vagotomized individuals from the Swedish National Patient Register during the period 1970-2020 and 40,855 non-vagotomized individuals matching for age, sex and type of peptic ulcer. The risk of being diagnosed with schizophrenia and associated psychoses (ICD10 codes F20-29) was analyzed using Cox proportional hazards regression models, including death as competing risk.

**Results:** When considering all types of vagotomy together, vagotomy was not significantly associated with schizophrenia (HR: 0.91 [0.72; 1.16]). However, truncal vagotomy (which denervates all subdiaphragmatic organs) significantly increased the risk of developing schizophrenia by 69% (HR: 1.69 [1.08; 2.64]), whereas selective vagotomy (which only denervates the stomach) showed no significant association (HR: 0.80 [0.61; 1.04]).

**Discussion:** Our results provide epidemiological support for the hypothesis that impairments in vagal functions could increase the risk of schizophrenia. Notably, the finding that truncal but not selective vagotomy is associated with an increased risk of schizophrenia raises the possibility that the activity of subdiaphragmatic non-gastric vagal branches may be of particular relevance for the development of schizophrenia.

## 1 Introduction

The vagus nerve is a major neuronal route of communication between the periphery and the brain ^1^. Vagal afferent neurons, the ascending component of this communication, sense peripheral signals and, thus, inform the brain about the state of the periphery. On the other hand, vagal efferent neurons provide top-down parasympathetic control over organ physiology.

Several lines of evidence suggest that this bidirectional communication is relevant for the development of schizophrenia. First, vagal parasympathetic activity is decreased in patients with schizophrenia, which contributes to overall autonomic nervous system dysfunction in affected individuals ^2^. This dysfunction underlies cardiovascular complications associated with schizophrenia, but is also increasingly postulated to contribute to psychological symptoms severity ^2^. In addition, vagal efferent neurons exert an anti-inflammatory action through the cholinergic anti-inflammatory pathway ^3^. Hence, impaired vagal function has been proposed to contribute to systemic as well as central inflammation seen in a subset of patients with schizophrenia ^4^.

In comparison to the descending vagal circuits, little focus has been given to ascending vagal afferents in schizophrenia. Several lines of evidence suggest, however, that vagal afferents may also play a significant role in pathophysiology. First, the activity of vagal afferent neurons directly influences the descending pathways and, hence, can contribute to the effects mentioned above. In addition, while recent studies emphasize the role of the gut microbiome in schizophrenia ^5, 6^, vagal sensory neurons are well-positioned to be the main mediators of the relevant microbiome metabolites ^7^. Finally, pre-clinical rodents studies indicate that vagal afferent signals could contribute to the development of schizophrenia-related abnormalities in brain and behavior. Indeed, subdiaphragmatic vagal deafferentation in rats leads to brain transcriptional changes in functional networks related to schizophrenia, increased sensitivity to dopamine-stimulating drugs, and impairments in sensorimotor gating and the attentional control of associative learning ^8^.

Despite these findings, direct evidence for a vagal influence on the pathophysiology of schizophrenia is lacking in humans. However, such evidence could foster further therapies targeted at the vagus nerve in schizophrenia and open avenues for more mechanistic studies.

Vagotomy, the lesion of the vagus nerve, was performed to treat peptic ulcers before the advent of more effective treatments, with the goal to reduce gastric acid secretion. Several vagotomy techniques co-existed and can be categorized into “truncal vagotomy”, the section of the vagal trunks below the diaphragm, and “selective vagotomy”, which aimed to selectively severe vagal sub-branches to the stomach ^9^.

Several epidemiological studies have taken advantage of cohorts of vagotomized individuals to assess vagal influence on dementia ^10^, Parkinson’s disease ^11^, inflammatory bowel disease ^12^ and mental disorders ^13^. While this last study included schizophrenia as a secondary outcome, a large study assessing the risk of developing schizophrenia in vagotomized individuals as a primary outcome is lacking.

Therefore, we built a nationwide retrospective cohort of patients who underwent vagotomy in Sweden and contrasted their risk of developing schizophrenia with that of control individuals matched for age, sex and peptic ulcer diagnosis over a 50-year period. In addition, we took advantage of the diverse vagotomy techniques to assess whether the extent of denervation modulates the risk of developing schizophrenia.

## 2 Methods

### 2.1 Ethics and reporting

This study was approved by the Swedish Ethical review authority (decision 2019-04833 and amendment 2021-05371-02). The current report is structured according to the STROBE guidelines (Strengthening the Reporting of Observational Studies in Epidemiology; ^14^).

### 2.2 Study design

This study is a register-based nationwide retrospective matched cohort covering the period 1970-2020 in Sweden.

### 2.3 Data sources

Anonymized data from different registers were provided by the Swedish National Board of Health and Welfare and Statistics Sweden. Specifically, the National Patient Register (NPR), the Swedish Cause of Death Register and the Total Population Register were used and linked by the Swedish personal identification number, which exists for each resident in Sweden ^15^. Since the early 1960s information from in-patients at public hospitals is documented in the NPR, including reports of psychiatric care. Nationwide coverage exists since 1987 ^16^. Furthermore, since 2001 outpatient information (doctor visits, psychiatric care and surgery) from private and public institutions are covered. Involuntary psychiatric care and forensic psychiatric care are also reported since 2008. Primary health care doctor visits are not yet documented in the NPR ^16^. The death register contains information on deaths from 1964. Regular, thorough quality and validity checks are done on data submitted to the National Board of Health and Welfare. Information about dates of immigration into (from 1969) and emigration out of (from 1961) Sweden is recorded in the Total Population Register.

### 2.4 Study population

A total of 49,170 individuals who underwent vagotomy between 01-01-1970 and 31-12-2020 and did not have a diagnosis of schizophrenia prior to surgery were identified from the National Patient Register based on the Swedish Classification of Operations and Major Procedures (Table S1) as described in ^12^. The closest diagnosis of a peptic ulcer (ICD-10 codes K25-K28, see Table S2) prior to vagotomy was considered as indication for vagotomy. Individuals identified from the NPR as vagotomized were excluded if surgery dates were not plausible (ie, before admission at the hospital or after death dates), if vagotomy surgery code was missing from the source file or if an ulcer diagnosis did not exist prior to or within 14 days post surgery. A maximum of 5 controls per case were identified from the NPR. All controls matching a vagotomy case for age at cohort entry, sex and presenting the same peptic ulcer diagnosis (exact ICD codes) were selected. The same control individuals could not be used as a control for multiple cases. We included all cases, irrelevant of the number of controls. Overall, 8,315 vagotomy patients and 40,855 controls, were included in the study, as described in the flow chart (Figure 1).

**Figure 1.**
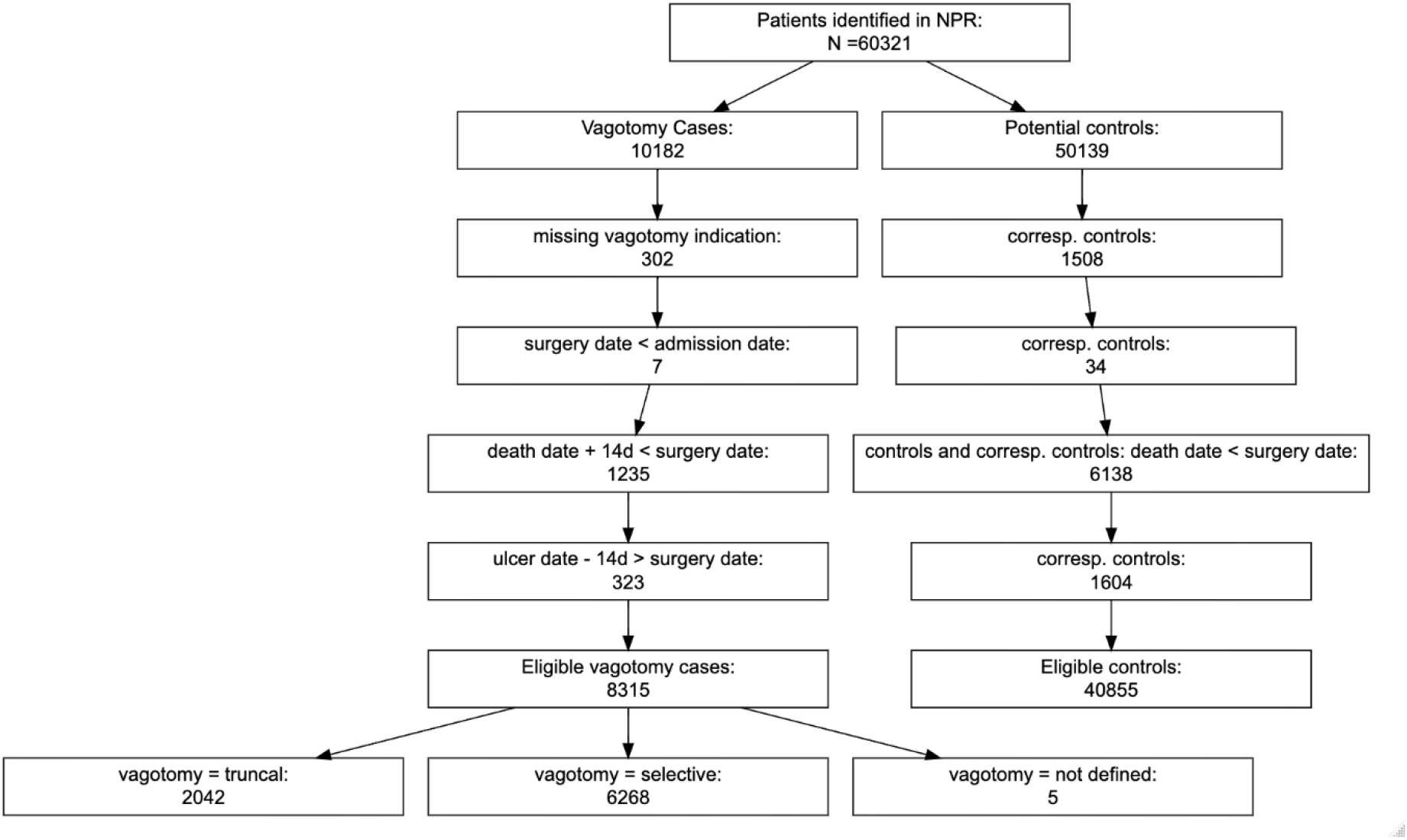
Flowchart. NPR = National Patient Registry

### 2.5 Ascertainment of surgery types

Vagotomy surgeries were categorized into “truncal” and “selective” as previously described in ^12^ based on the Swedish classification of Operations and Major procedures (Table S1). Briefly, truncal vagotomy lesions both vagal trunks around the oesophagus and therefore denervate all subdiaphragmatic organs. Selective vagotomies cover multiple techniques achieving a selective denervation of the stomach ^17^. Five individuals had surgery codes that cannot be assigned to truncal or selective and were therefore only considered for the analysis of the overall vagotomy effects.

### 2.6 Ascertainment of schizophrenia diagnosis

Due to historical evolution in the diagnosis of schizophrenia ^18^ and the heterogeneity of the disease ^19^, we opted for a broad definition of schizophrenia for this long-term nationwide cohort. Hence, diagnoses of schizophrenia, schizotypal disorders and non-mood psychotic disorders were identified by the presence of the corresponding ICD codes in the NPR or the Death register (ICD10 codes F20-F29; see Table S3 for earlier ICD versions). We refer to these disorders as “schizophrenia” in the rest of the manuscript. In case of several diagnoses, the earliest diagnosis and diagnosis dates were considered. Importantly, diagnoses of schizophrenia as reported in the Swedish NPR match to a high degree with individual medical records of schizophrenia diagnoses according to DSM criteria ^20^.

### 2.7 Covariates

Age and sex of participants were taken from the NPR. Overall comorbidity at cohort entry was assessed with an adapted version ^21^ of the weighted Charlson Comorbidity Index (CCI) ^22, 23^. The CCI is a cumulative score including diagnoses of vascular diseases, diabetes, chronic obstructive pulmonary disease, mild, moderate and severe liver disease, cancer and chronic obesity. Relevant ICD codes can be found in supplementary material (Table S4).

### 2.8 Handling of missing data

For 2,104 patients in the death registry, exact day of death was not indicated and was imputed as the 15^th^ of the corresponding month. Further, there were 410 cases for which month and day of death were missing. In this case, we used December 15^th^ of the respective year of death.

### 2.9 Statistical methods

Cohort participants were followed up from the day of surgery, which was matched for the reference population, to the date of a schizophrenia diagnosis, death, emigration of Sweden or December 31^st^, 2020, whichever occurred first.

The first analysis assessed the association between overall vagotomy and the postoperative risk of developing schizophrenia. In a secondary analysis, the association between vagotomy types (truncal, selective) and schizophrenia was investigated. In a third analysis, we restricted follow up to 5, 10, 15 and 20 years after cohort entry and analyzed the association between overall vagotomy, as well as vagotomy types (truncal, selective) and schizophrenia.

For all analysis, Cox proportional hazard regression models and hazard ratios with corresponding 95% confidence intervals were computed for the outcome schizophrenia, including death as a competing risks. We considered a crude model, accounting for matching variables age and sex at cohort entry, and for the number of controls per case, as well as an adjusted model, additionally including the ICD period and the adapted Charlson Comorbidity Index. All analyses were conducted with R (version 4.3.1).

## 3 Results

Table 1 summarizes characteristics of the study participants. As a result of matching, the distributions of sex (66% males), age (median age 54 for controls, median age 53 in the vagotomy group) and peptic ulcer types were similar in vagotomized patients and controls. Most vagotomy patients (n = 6,268, 75.4 %), underwent selective vagotomy. Individuals with truncal vagotomy had higher comorbidity (CCI) at baseline and higher mortality during follow-up than reference individuals or individuals with selective vagotomy.

**Table 1.**
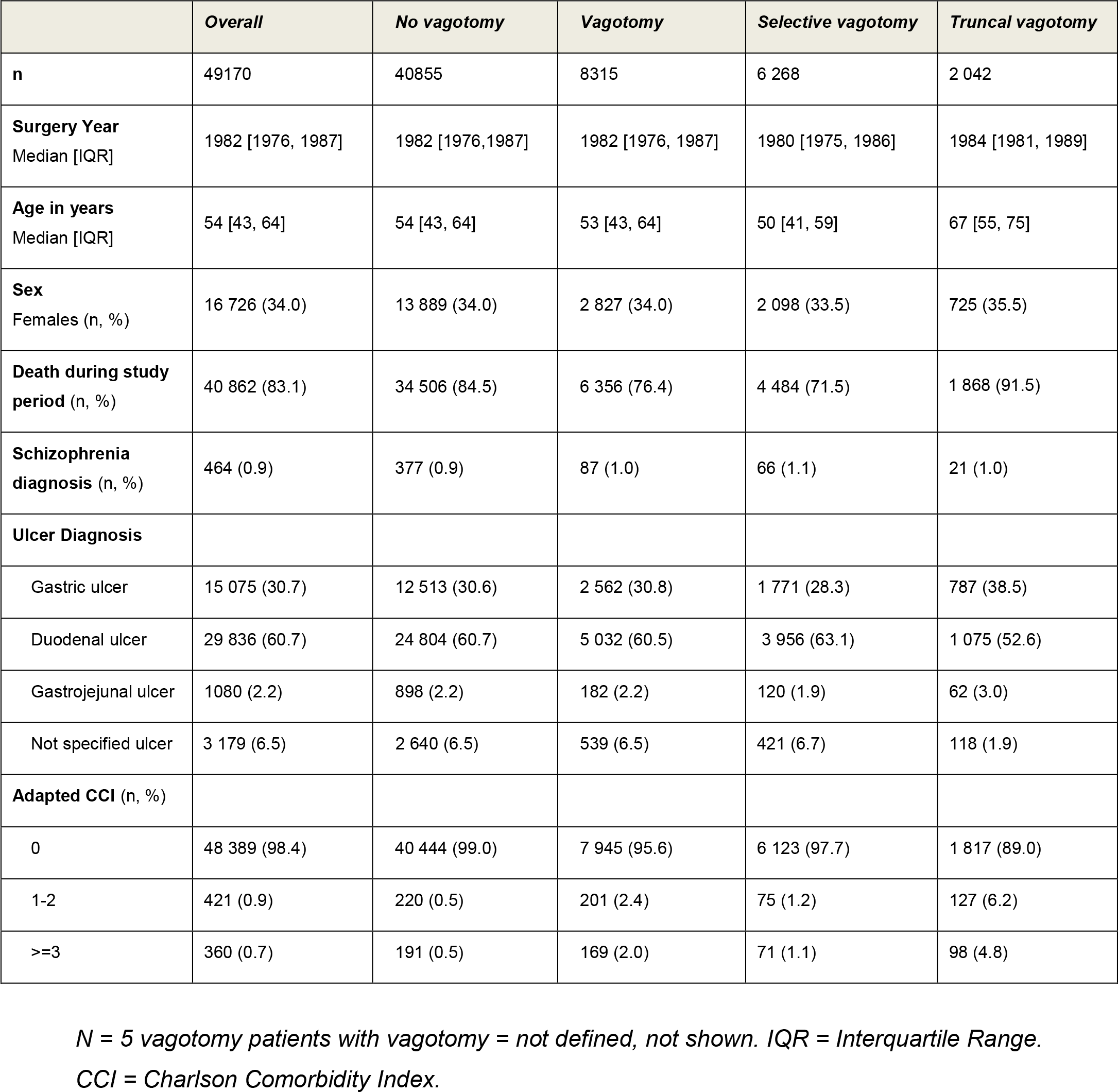
Patient characteristics.

The cumulative incidence of schizophrenia in vagotomized patients compared to the matched reference individuals is shown in Figure 2 (overall vagotomy) and Figure 3 (truncal and selective vagotomy). Overall, we found no statistically significant association of vagotomy with schizophrenia (HR crude model: 0.93 [0.73; 1.18]; adjusted 0.91 [0.72; 1.16], Figure 4). When considering vagotomy types, however, truncal vagotomy increased the subsequent risk of schizophrenia (HR adjusted model: 1.69 [1.08; 2.64]), while selective vagotomy show no significant association (HR adjusted model: 0.80 [0.61; 1.04], Figure 5). In all analyses, the effect of vagotomy did not differ by sex, as shown by the absence of significant vagotomy*sex interactions. (Table S5).

**Figure 2.**
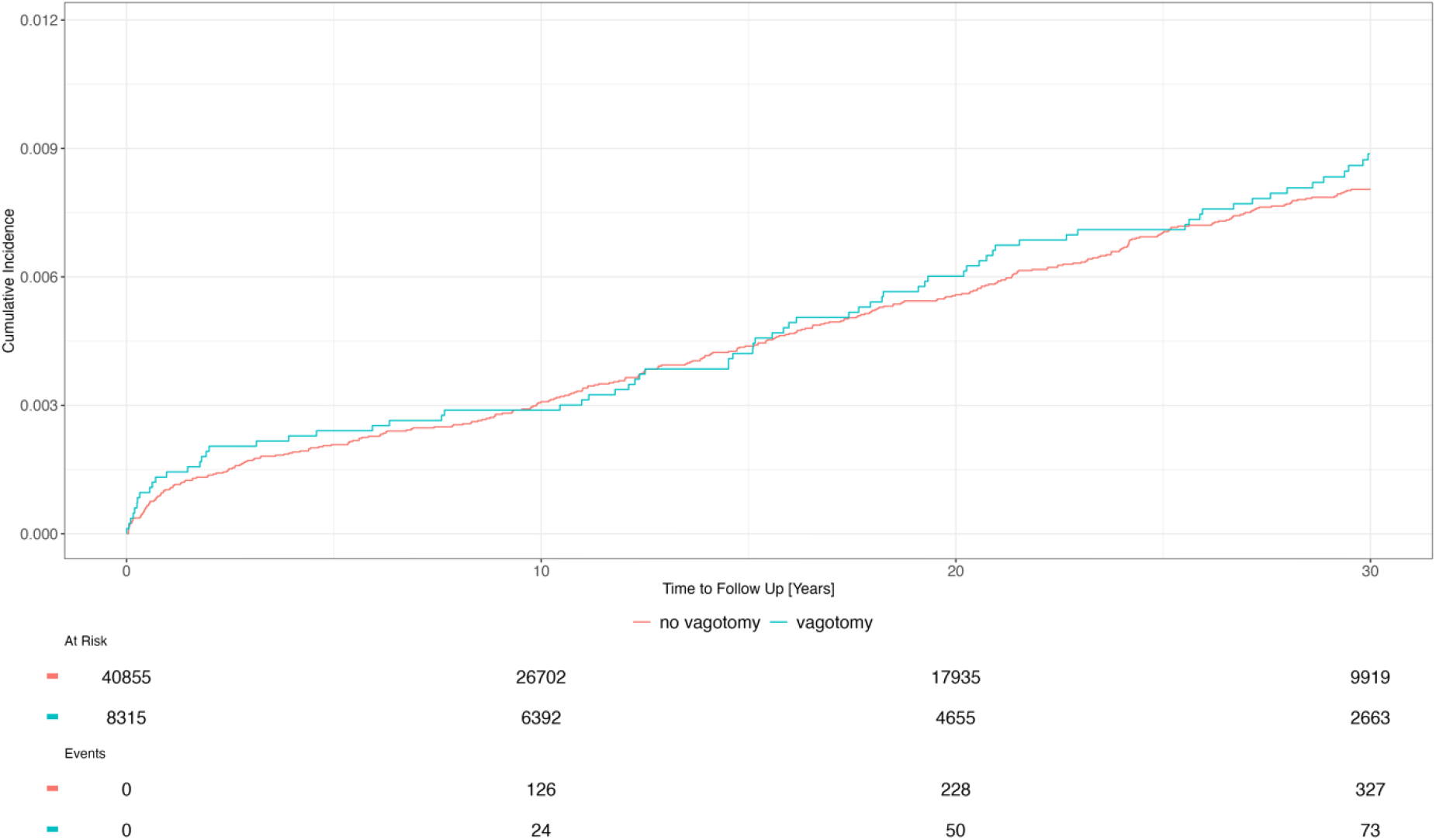
Cumulative incidence function of schizophrenia among patients with overall vagotomy and matched controls.

**Figure 3.**
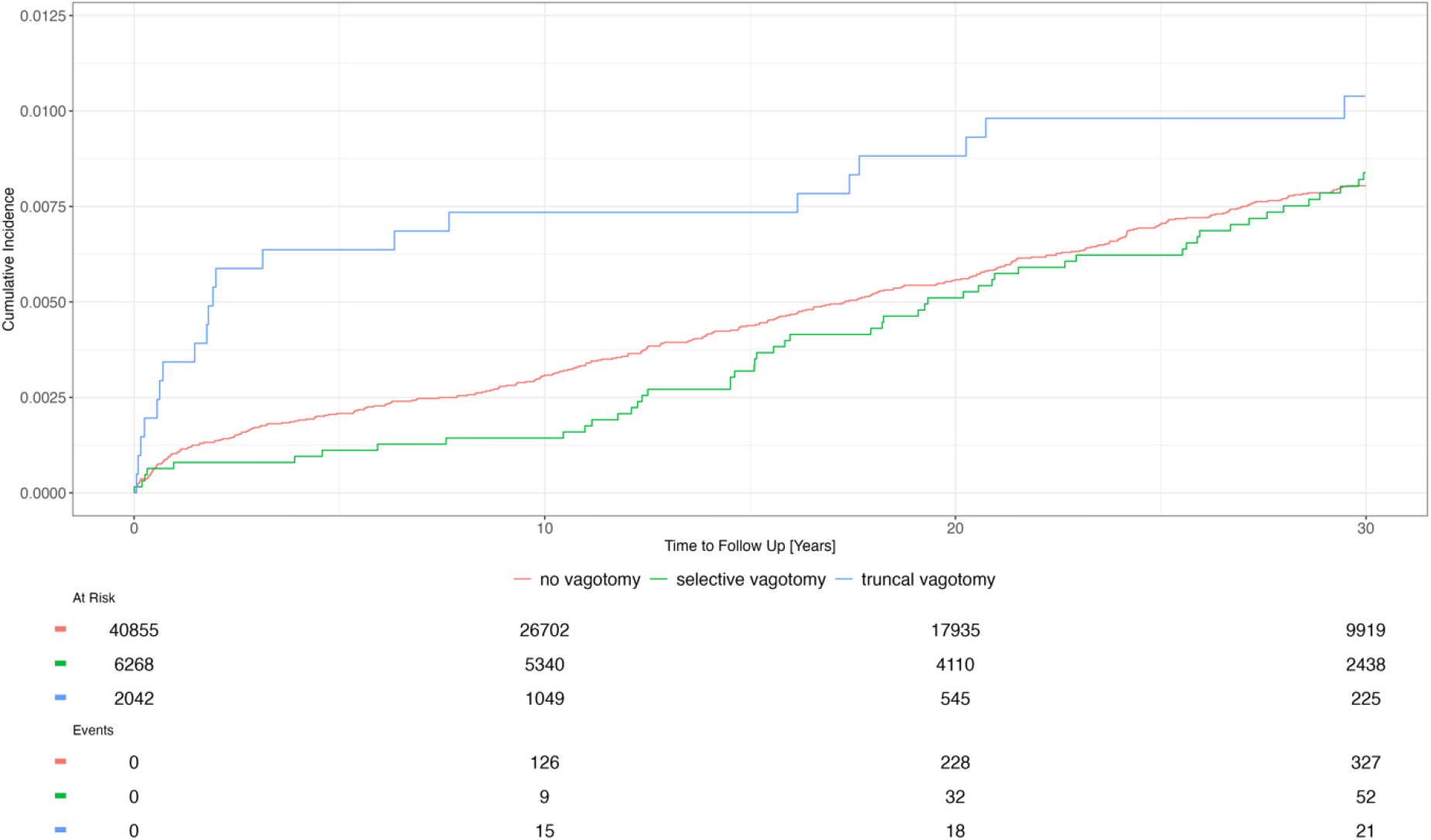
Cumulative incidence function of schizophrenia among patients with truncal or selective vagotomy and matched controls.

**Figure 4.**
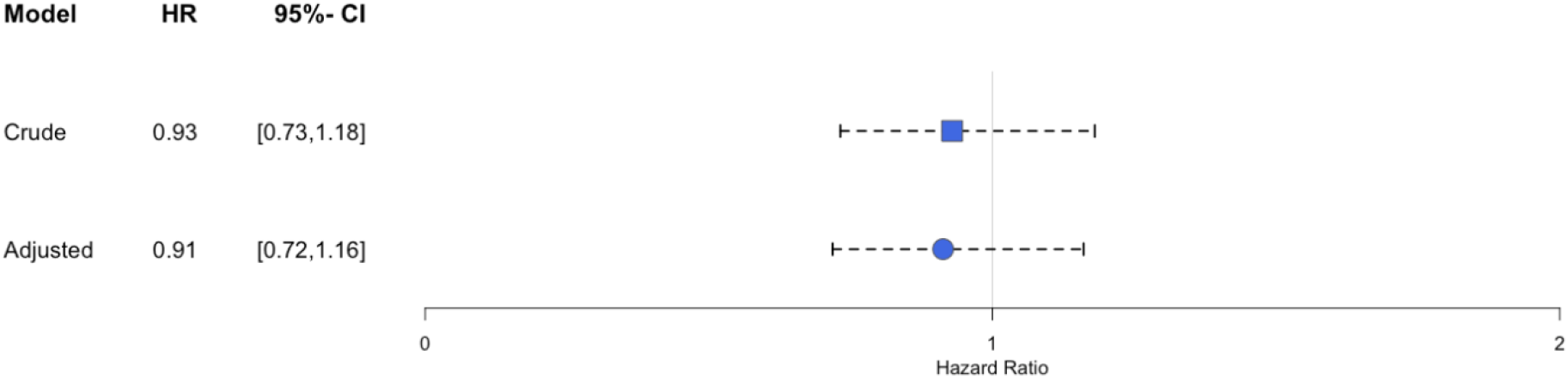
Regression Results of Cox proportional hazards model for overall vagotomy and subsequent diagnosis of schizophrenia. Crude model controlled for age, sex, and controls per case. Adjusted model controlled for age, sex, weighted Charlson Comorbidity Index, ICD-period, controls per case. HR = Hazard Ratio, 95% CI = 95% Confidence Interval. Vertical line: reference value.of 1

**Figure 5.**
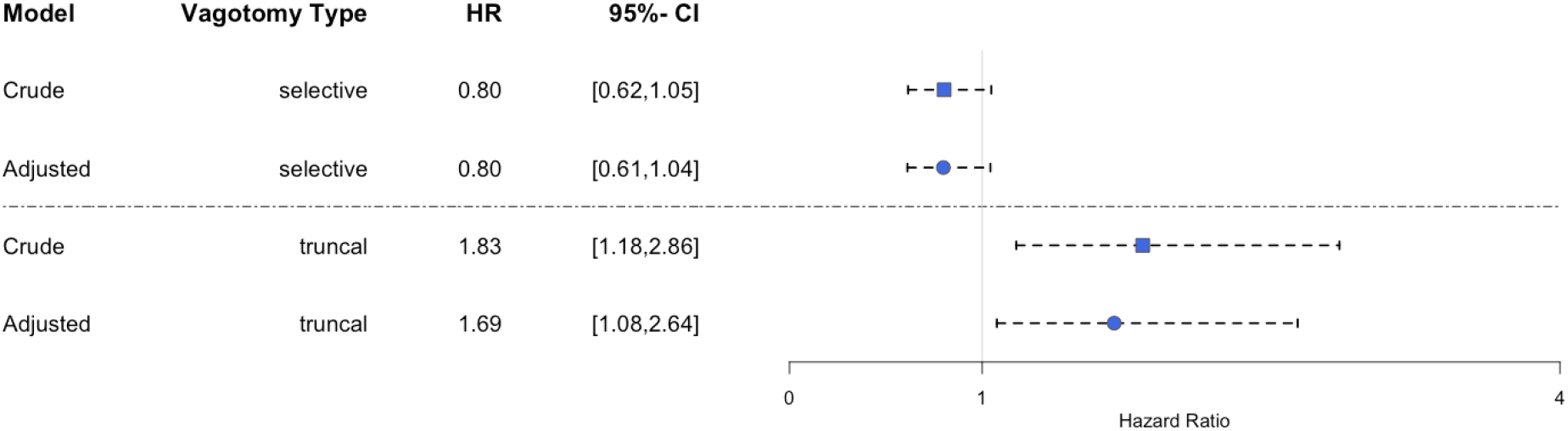
Regression results of Cox proportional hazards model for selective or truncal vagotomy and subsequent diagnosis of schizophrenia. Crude model controlled for age, sex, and controls per case. Adjusted model controlled for age, sex, weighted Charlson Comorbidity Index, ICD-period, controls per case. HR = Hazard Ratio, 95% CI = 95% Confidence Interval. Vertical line: reference value of 1

When the temporal relationship of vagotomy and schizophrenia was analysed by restricting follow up to 5, 10, 15 and 20 years, we found no statistically significant effect of overall vagotomy at any time point (Table S6). The positive association between truncal vagotomy and schizophrenia was statistically significant at all times (Figure 6 and Table S7). The effect was highest after 5 years (adjusted HR: 4.03 [2.21, 7.34]) and the lowest after 15 years (15 years adjusted HR: 2.04 [1.18, 3.54]). Selective vagotomy showed a protective effect on the development of schizophrenia at 10 and 15 years (Figure 6 and Table S7).

**Figure 6:**
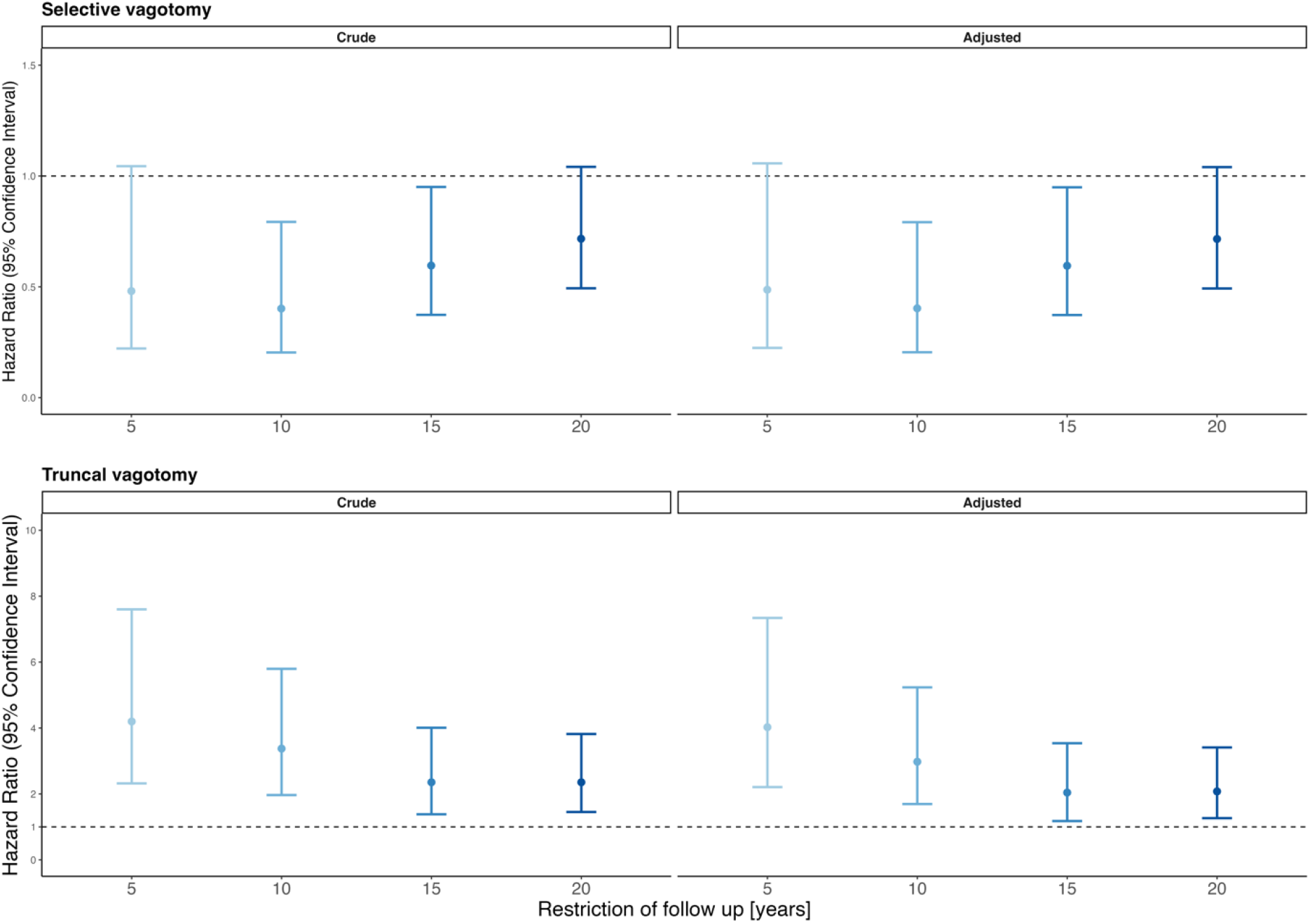
Regression results of Cox proportional hazards model for selective or truncal vagotomy and schizophrenia, with follow-up restriction. Crude model controlled for age, sex, and controls per case. Adjusted model controlled for age, sex, weighted Charlson Comorbidity Index, ICD-period, controls per case. HR = Hazard Ratio, 95% CI = 95% Confidence Interval. Vertical line: reference value of 1

## 4 Discussion

We built a nationwide retrospective cohort of individuals who underwent vagotomy and contrasted their risk of developing schizophrenia to that of control individuals. Our study included a total of 49,170 individuals identified in the Swedish NPR over a 50-year period. We used a novel approach where controls are matched with vagotomized patients for age, sex and peptic ulcer diagnosis, enabling us to correct for confounding by indication, a challenge faced by previous studies ^10–13^.

An earlier study assessed the risk of developing mental disorders in a nationwide sample of vagotomized individuals in Denmark, and included the risk of developing schizophrenia as a secondary outcome ^13^. With 38 cases identified in the vagotomy group, this study observed no effect of vagotomy on schizophrenia, although with large confidence intervals. Here, we leveraged a larger national register, started the follow-up at an earlier date (which is important given the temporal window in which vagotomies were performed) and used a longer study period. These modifications in the study design yielded more observations and cases than the Danish study (87 in the vagotomy group, 464 overall). Nevertheless, in accordance with this study, we found no effect of vagotomy overall on the subsequent risk of developing schizophrenia.

Vagotomy, however, covers several techniques ^17^: truncal vagotomies severe both vagal trunks at the level of the oesophagus and hence, remove vagal innervation from all subdiaphragmatic organs (*ie*, gastrointestinal tract, liver, pancreas …). In contrast, selective vagotomies only interrupt vagal innervation from the stomach. Our study is the first to distinguish between the two types of vagotomies for the outcome of schizophrenia. Intriguingly, we found that truncal vagotomy significantly increases, while selective vagotomy was not associated with the risk of developing schizophrenia compared to matched controls. Interestingly, Bunyoz et al. also found that truncal vagotomy, but not selective vagotomy, was associated with an increase in mental disorders ^13^.

A first potential explanation of these differences may lie in the distinct indication between selective and truncal vagotomy and a possible surveillance bias. Selective vagotomy is a more subtle lesion with less side effects, but is associated with a higher recurrence rate than truncal vagotomy. Hence, truncal vagotomy was primarily performed in patients where recurrence rate needed to be minimized despite side effects, such as elderly patients or patients with comorbidities ^24^. It has been argued that these patients are more likely to visit the hospital after surgery than individuals with selective vagotomy and this could explain the different effects of vagotomy types on the development of schizophrenia (surveillance bias) ^13^. Here, we matched controls for age, sex and ulcer diagnosis, which mitigates this bias. Moreover, the effect of truncal vagotomy on other outcomes (inflammatory bowel disease, dementia) tends to be similar to that of selective vagotomy ^10, 12^. Truncal vagotomy even reduces the risk of developing Parkinson’s disease, while selective vagotomy had no effect ^11^. Hence, the surveillance bias explanation seems unlikely and the observed differences might be rooted in biology.

The findings that truncal, but not selective vagotomies, are associated with an increase in the risk of developing schizophrenia indicate a specific role of non-gastric subdiaphragmatic branches of the vagus nerve in the aetiology of schizophrenia. Among the non-gastric vagal branches are notably those that project to the intestine. It is therefore tempting to hypothesize that intestinal signals, such as those induced by shifts in the intestinal microbiome ^7^, could influence schizophrenia-relevant brain circuits via gut-projecting vagal sensory neurons. Novel branch-selective vagal manipulation techniques in rodent models could address this hypothesis ^25, 26^. Moreover, beyond the concept of anatomically-defined branches, a rapidly-expanding field is now unravelling the molecular and functional diversity of vagal afferent and efferent neurons ^27–29^. Recent rodent findings have confirmed the notion that molecularly-defined vagal neuronal populations have distinct functions, even when innervating the same organs: for example, only a subset of vagal sensory neurons projecting throughout the gastrointestinal tract contributes to eating behavior, or reward behavior modulation ^27, 30^. Distinct vagal populations can even have opposing actions, as exemplified by two vagal sensory subtypes that induce apnea or rapid breathing ^31^. Despite this emerging granularity in our understanding of vagal circuits, whether specific molecularly-defined subdiaphragmatic vagal neurons are of particular relevance for the risk of schizophrenia remains unknown. This opens avenues for future studies evaluating the role of distinct neuronal populations on schizophrenia-like behaviors, for example with the help of genetically-engineered mouse models.

Vagal neurons, specifically vagal afferent neurons, are well positioned to modulate central pathways involved in the pathophysiology of schizophrenia. In the rat, a vagal lesion that primarily affects subdiaphragmatic vagal afferent neurons, was associated with brain transcriptomic changes annotating with schizophrenia-associated disease pathways and increased striatal dopamine content ^8^. Interestingly, increased dopamine content within striatal structures is a core pathophysiological mechanism in schizophrenia ^32^. It is thought to underlie the disruption of attentional control of associative learning ^33^, sensorimotor gating ^34–36^, and amphetamine hypersensitivity ^37^. From a circuit perspective, ascending vagal inputs to the nucleus of the solitary tract are synaptically connected to the ventral tegmental area ^30^, which contains the majority of dopaminergic cells projecting to the ventral striatum. Beyond mesolimbic dopamine, future studies should examine the contribution of vagal neurons to schizophrenia-relevant brain circuits in a more systematic manner.

Finally, the relevance of our findings extends beyond the specific case of vagotomized individuals. Reduced parasympathetic activity, evidenced by a low heart rate variability, is a common feature of many mental and metabolic disorders, suggesting that descending vagal activity is also affected in a large population of patients ^38^. In addition, vagal afferent neurons show reduced response to gastrointestinal hormones or stretch in animal models of obesity ^39–42^ or animals with microbial dysbiosis ^43^. Hence, if these findings extend to humans, the number of individuals affected by impairments in vagal ascending pathways is likely to be large.

Overall, our study confirms the absence of effect of overall vagotomy on schizophrenia seen in a previous study. It opens, however, the possibility that distinct vagal circuits play a differential role in the risk of developing schizophrenia. Notably, our findings suggest a role for the non-gastric subdiaphragmatic branches of the vagus nerve in the development of schizophrenia. While future epidemiological studies are warranted to strengthen this conclusion, studies in rodents are well-suited to test this hypothesis and uncover the underlying mechanisms.

## 7 Author contributions

CFR: Methodology, Formal analysis, Investigation, Visualization, Writing-Original draft preparation

KPS: Conceptualization, Writing-Reviewing and Editing

SR: Conceptualization, Writing-Reviewing and Editing

UM: Writing-Reviewing and Editing

JPK: Conceptualization, Methodology, Writing-Original draft preparation, Writing-Reviewing and Editing

## Data Availability

All aggregated data produced in the present study are available upon reasonable request to the authors. Individual data need to be requested to competent registry.

