## Supplementary data for "A vagal influence on schizophrenia? A nationwide retrospective cohort of vagotomized individuals"

**Affiliations** :
1 Institute of Veterinary Pharmacology and Toxicology, University of Zurich-Vetsuisse, Switzerland
2 Institute of Neuroscience and Physiology, University of Gothenburg, Sweden
3 Department of Nutritional Sciences, Pennsylvania State University, State College, PA, United States
4 Neuroscience Center Zurich, University of Zurich and ETH Zurich, Switzerland
5 Division of Chronic Disease Epidemiology, Epidemiology, Biostatistics and Prevention Institute, University of Zurich, Switzerland

**Table S1: Surgery codes used for inclusion of vagotomized individuals and categorization into "selective" and "truncal" vagotomy**

| | Years | Swedish Classification of Operations and Major Procedures Codes |
|---|---|---|
| **Selective** | 1964 - 1996 | 4470, 4474, 4475, 4476, 4477, 441, 4414, 4416 ,4419 |
| | 1997 - | JDG10, JDG11 |
| **Truncal** | 1964 - 1996 | 4471, 4472, 4473, 4478, 4411, 4413, 4415, 4418, 4451, 4453 |
| | 1997 - | JDG00, JDG01 |
| **Non-defined** | 1964 – 1996 | - |
| | 1997 - | JDG96, JDG97 |

**Table S2: ICD codes considered for peptic ulcers**

| | ICD8 (1969-1986) | ICD9 (1987-1996) | ICD10 (1997- ) |
|---|---|---|---|
| **Gastric ulcer** | 531 | 531 | K25 |
| **Duodenal ulcer** | 532 | 532 | K26 |
| **Non-specified** | 533 | 533 | K27 |
| **Gastrojejunal ulcer** | 534 | 534 | K28 |

**Table S3: ICD codes considered for the main outcome "schizophrenia"**

| | ICD7 ( - 1968) | ICD8 (1969-1986) | ICD9 (1987-1996) | ICD10 (1997- ) |
|---|---|---|---|---|
| **Schizophrenia, schizotypal disorder and non-mood psychotic disorders** | 300 | 295 | 295 | F20-F29 |

**Table S4: ICD codes considered for the adapted Charlson Comorbidity Index**

*COPD = Chronic obstructive pulmonary disease, CVD = Cerebrovascular Disease, MI = Myocardial Infarction, PVD = Peripheral Vascular Disease, CHD = Congestive Heart Failure*

| | ICD7 ( - 1968) | ICD8 (1969-1986) | ICD9 (1987-1996) | ICD10 (1997- ) |
|---|---|---|---|---|
| **COPD** | 502 | 491, 492 | 491, 492, 496 | J41, J42, J43, J44 |
| **Obesity** | 278 | 277 | 278 | E66 |
| **Diabetes** | 260 | 250 | 250 | E10, E11, E12, E13, E14 |
| **Liver disease** | 581.1 | 571.0 | 571A, 571B, 571C, 571D | K70 |
| **Cancer** | 140-199, 200-205 | 140-149, 150-159, 160-163, 180-189, 190-199, 200-209 | 140-149, 150-159, 160-165, 170-175, 179-189, 190-199, 200-208 | Cxx |
| **MI** | 420.1, 420.9 | 410 | 410, 412 | I21, I22, I25.2 |
| **PVD** | 450, 451, 453.3, 455 | 411, 443.9, 445 | 441, 443X, 785E | 1997 I70, I71, I73.1, I73.8, I73.9, I77.1, I79.0, 79.2, K55.1, K55.8, K55.9, Z95.8, Z95.9 |
| **CVD** | 330-334 | 430-438 | 430-438 | G45, G46, H34.0, I60-I69 |

**Table S5: Regression results of Cox proportional hazards model for interaction of vagotomy*sex and schizophrenia.**

| | Crude | | Adjusted | |
|---|---|---|---|---|
| | **HR** | **95% - CI** | **HR** | **95% - CI** |
| | **Adjusted for Overall Vagotomy** | | | |
| **Vagotomy, Sex = Female** | 1.07 | [0.67, 1.70] | 1.07 | [0.67, 1.71] |
| | **Adjusted for Vagotomy Types** | | | |
| **Selective, Sex = Female** | 1.23 | [0.73, 2.08] | 1.24 | [0.74, 2.09] |
| **Truncal, Sex = female** | 0.67 | [0.26, 1.71] | 0.67 | [0.26, 1.69] |

*Crude model controlled age and interaction term vagotomy and sex, controls per case. Adjusted model controlled for age, interaction term vagotomy and sex, weighted Charlson Comorbidity Index, ICD period, controls per case. HR = Hazard Ratio, 95% CI = 95% Confidence Interval.*

**Table S6: Regression results of Cox proportional hazards model for overall vagotomy and schizophrenia, with follow-up restriction.**

| | Crude | | Adjusted | |
|---|---|---|---|---|
| | HR | 95% - CI | HR | 95% - CI |
| **5 years** | 1.13 | [0.69, 1.86] | 1.12 | [0.68, 1.83] |
| **10 years** | 0.89 | [0.58, 1.39] | 0.86 | [0.55, 1.34] |
| **15 years** | 0.88 | [0.61, 1.27] | 0.85 | [0.58, 1.23] |
| **20 years** | 0.96 | [0.70, 1.30] | 0.93 | [0.68, 1.27] |

*Crude model controlled age, sex, controls per case. Adjusted model controlled for age, sex, weighted Charlson Comorbidity Index, ICD period, controls per case. HR = Hazard Ratio, 95% CI = 95% Confidence Interval.*

**Table S7: Regression results of Cox proportional hazards model for selective or truncal vagotomy and schizophrenia, with follow-up restriction.**

| | Crude | | Adjusted | |
|---|---|---|---|---|
| | HR | 95% - CI | HR | 95% - CI |
| | | **Selective vagotomy** | | |
| **5 years** | 0.48 | [0.22, 1.04] | 0.49 | [0.22, 1.06] |
| **10 years** | 0.40 | [0.20, 0.79] | 0.40 | [0.20, 0.79] |
| **15 years** | 0.60 | [0.37, 0.95] | 0.59 | [0.37, 0.95] |
| **20 years** | 0.72 | [0.49, 1.02] | 0.72 | [0.49, 1.04] |
| | | **Truncal vagotomy** | | |
| **5 years** | 4.20 | [2.32, 7.60] | 4.03 | [2.21, 7.34] |
| **10 years** | 3.38 | [1.97, 5.80] | 2.98 | [1.69, 5.23] |
| **15 years** | 2.35 | [1.38, 4.01] | 2.04 | [1.18, 3.54] |
| **20 years** | 2.35 | [1.45, 3.82] | 2.08 | [1.26, 3.41] |

*Crude model controlled age, sex, controls per case. Adjusted model controlled for age, sex, weighted Charlson Comorbidity Index, ICD period, controls per case. HR = Hazard Ratio, 95% CI = 95% Confidence Interval.*
